# Sentinel Coronavirus Environmental Monitoring Can Contribute to Detecting Asymptomatic SARS-CoV-2 Virus Spreaders and Can Verify Effectiveness of Workplace COVID-19 Controls

**DOI:** 10.1101/2020.06.24.20131185

**Authors:** Douglas L. Marshall, Frederic Bois, Soren K. S. Jensen, Svend Aage Linde, Richard Higby, Yvoine Rémy-McCort, Sean Murray, Bryan Dieckelman, Fitri Sudradjat, Gilles G. Martin

**Author notes:** **Address for Correspondence:** Douglas Marshall, Eurofins Microbiology Laboratories, 2000 Mackenzie Ct., Fort Collins, CO 80528 USA; 1-970-217-6854.

## Abstract

Detecting all workplace asymptomatic COVID-19 virus spreaders would require daily testing of employees, which is not practical. Over a two week period, nine workplace locations were chosen to test employees for SARS-CoV-2 infection (841 tests) and high-frequency-touch point environmental surfaces (5,500 tests) for Coronavirus using Eurofins COVID-19 Sentinel^™^ RT-PCR methods. Of the 9 locations, 3 had one or employees infected with SARS-CoV-2, neither of whom had symptoms at the time of testing nor developed symptoms. Locations with Coronavirus contaminated surfaces were 10 times more likely to have clinically positive employees than locations with no or very few positive surfaces. Break room chairs, workbenches, and door handles were the most frequently contaminated surfaces. Coronavirus RNA was detected at very low concentrations (RT-PCR 34 to 38 C_q_). Environmental monitoring can be used to validate intervention strategies and be useful to verify the effectiveness of such strategies on a regular basis.

## Introduction

Airborne Coronavirus shedding into the environment is a known vehicle of transmission (*1*). Depending on viral load in expelled coughs, sneezes, and other exhalations, air currents, and proximity to active shedder, broad workplace or social spread of this disease can occur in confined spaces (*2*). Once deposited on surfaces, this viral load can persist for hours or days (*3*) can serve as a fomite to transmit respiratory pathogens, such as Coronavirus. Modeling has estimated the majority of COVID-19 outbreak transmission occurs due to exposure to presymptomatic individuals (before showing symptoms) (46%), asymptomatic individuals (who never show symptoms) (10%), and contaminated environmental sources (6%), while transmission by exposure to symptomatic individuals accounts for 38% of cases (*4*). Others have shown that exposure to presymptomatic individuals up to 5 days before symptoms was the leading cause of infections compared to exposure to symptomatic individuals (*5, 6*).

In response to the COVID-19 pandemic, governments and employers have deployed a set of common measures to control spread of the SARS-CoV-2 virus in workplace and social environments, including frequent disinfection, fitness for duty screens (temperature checks, symptom checklist, and exposure history to symptomatic individuals), social distancing, staggered work shifts, small team segregation, and work from home policies. Unfortunately, such actions have been either poorly implemented or are not of sufficient efficacy to fully prevent workplace COVID-19 outbreaks. The primary reason for workplace control failure is the inability to detect pre- and asymptomatic carriers (*7*). Such carriers can shed virus in the work environment and expose fellow workers to a steady stream of virus leading to air and fomite transmission spread (*4, 8, 9, 10, 11, 12*).

Individuals who are actively presenting with COVID-19 symptoms likely will stay home from work and not attend social events; however, asymptomatic, presymptomatic, or mildly symptomatic SARS-CoV-2 infected individuals may report for work and socialize. In addition, some workplace cultures have a “work while sick” policy (never written), those deciding not to work are not given paid sick leave, resulting in economic penalty. Those that work in a critical industry also may be motivated to work when ill. Due to perceived shortage in testing capabilities in many regions, such individuals also may be denied testing for SARS-CoV-2. An as yet unresolved problem seems to be to find a way to detect asymptomatic or presymptomatic COVID-19 carriers that does not require testing each employee every day.

Employee testing practices and associated social challenges are proving difficult due to inconvenience, expense, invasiveness, and sometimes painful experiences. In many jurisdictions, employers may not have legal access to employee test results due to privacy laws. In others, employee unions may disallow such testing or results access. And perhaps most importantly, systematic worker antibody testing may raise discrimination questions: which workers are discriminated against – those who have had exposure to the virus or those that haven’t? An ideal situation for business continuity is to have a way to early detect employees who may be active virus spreaders without symptoms and to detect employees who may be convalescing from symptomatic COVID-19 illness and beyond a recommended quarantine window, yet still shedding virus before any of those can infect others in the workplace.

It is possible to swab every day (or every evening), each room of a workplace building and if the virus is detected to quarantine and test the employees who were present in that room. Alternatively in cities where the virus prevalence is very low, sentinel testing can be done on high-frequency touch-point surfaces in common areas or wastewater plants (*13, 14*). The objectives of this study were to assess whether Coronavirus environmental monitoring has use as a tool to detect asymptomatic and presymptomatic virus spreaders and to assess effectiveness of workplace COVID-19 management plans by conducting clinical and environmental surface testing at work locations.

## Materials and Methods

### Study Location Selection

Nine workplace locations in Europe and the United States of America were voluntarily enrolled in the study based on evidence of community COVID-19 occurrence. All locations had written COVID-19 control plans prior to study initiation. Location facilities were small to mid-sized urban/suburban class A or B office or mixed-use industrial buildings. Europe locations were Brussels Belgium, Madrid Spain, Milano Italy, Saverne France, and Wolverhampton United Kingdom. United States locations were Battle Creek Michigan, New Brunswick New Jersey, New Orleans Louisiana, and Portage Michigan. A volunteer study manager was appointed at each location to manage administrative tasks such as sampling logistics, employee testing paperwork at the site, and to ensure respect of privacy and healthcare laws. Managers were trained on the study protocol and sampling techniques.

At the end of the study a tenth location was enrolled. This location was a nonprofit faith-based organization serving seniors older than 53 years with 600 staff and 3,000 residents located near St. Cloud Minnesota USA. The organization offers independent and assisted living, transitional, long term and memory care services among others. Some of these residents were clinically confirmed COVID-19 positive. We were provided an opportunity to swab environmental surfaces in one infected resident’s room and adjacent employee work areas.

### Employee SARS-CoV-2 Testing

Most study locations had two rounds of employee clinical testing during week 1 and 2 of the study. The Saverne location had an addition round during week 3 and the St. Cloud location did not participate in clinical testing. Employees at the testing locations were asked to volunteer for the study. To be eligible, employees must have been physically working at the study location prior to the study launch. Work-from-home employees were excluded. Employees were neither paid nor coerced to volunteer for the study. Consent forms appropriate for the local regulatory jurisdiction were processed for each employee prior to testing. Eligible employee participation rate ranged from 56 to 100%. Employees were administered either nasopharyngeal swabs (iClean® Nasopharyngeal Nylon Flocked Swab, Chenyanglobal Group, Shenzhen, China) at Europe locations or oropharyngeal swabs (Opti-Swab, Puritan Medical Products Company, Guilford, Maine, USA) at USA locations by a qualified health-care specialist.

After collection, labeled swabs were sent via overnight courier to accredited clinical testing laboratories (Eurofins LifeCodexx, Konstanz, Germany for Europe locations or Viracor Eurofins Clinical Diagnostics, St. Charles, Missouri for USA locations) and tested for presence of SARS-CoV-2 using multiplex RT-PCR assays for the envelop E gene RNA (RIDA®GENE SARS-CoV-2 RUO, r-biopharm, Darmstadt, Germany) for Europe locations or nucleocapid N gene RNA (Viracor April 6, 2020 U.S. Food and Drug Administration Emergency Use Authorization EUA200124) for USA locations. PCR reactions with C_t_ values ≤38 were considered detected for SARS-CoV-2 RNA, with an LOD of 73 copies/mL

### Environmental Swab Testing

To establish coronavirus baseline surface contamination rate, 2 days of preliminary testing was conducted at the New Orleans study location prior to clinical testing. On both days, 24 high-frequency-touch point surfaces were swabbed, resulting in 48 samples. Thereafter, each study location collected daily environmental swabs on days 1 through 14 of the study, with no swabbing conducted on weekend days. A list of high-frequency-touch point surfaces was provided to each study location following Eurofins Sentinel^™^ Program guidelines (Table 1), but other touch-point surfaces could be chosen for sampling. The study manager at each location risk ranked each sampling surface based on frequency of touching and the number of people who contact such surfaces during a normal work day. Greater risk surfaces were those that were touched frequently by a large number of the workforce or received infrequent cleaning and disinfection.

**Table 1.** Recommended high-frequency-touch-point sampling sites – not in order of priority.

|  |
| --- |
| Floors |
| Door handles, knobs, & push plates |
| Walkway, stairway hand rails |
| Copy machine touch pad and cover |
| Shared keyboards, mouse, I pads, phones, headsets, PDAs & remote controllers |
| Time clocks |
| Door/gate/elevator keypads & buttons |
| Light switches |
| Table/bench/desk tops near seating area |
| Drawer/cabinet knobs |
| Chair back tops, armrests & seat sides |
| Sink faucet handles/nobs |
| Cleaning supplies – broom/mop handles, spray bottles, soap/sanitizer dispensers if not automatic, & towel dispensers if not automatic |
| Forced air hand dryers |
| Trash bin rims & lids |
| Appliance/equipment control panels, touch pads, control nobs & handles |
| Coffee dispensers |
| Utensils & tools |
| Vending machine touch pad & product dispenser chute |
| Toilet flush handles, nobs & seats |
| Toilet room stalls doors & latches |
| Feminine napkin dispensers |
| PPE storage bins |
| Employee lockers |
| Log books |
| Air handling unit vents & filters |

On each sampling day, a total of 15 surface swabbing surfaces were selected based on the study location risk ranking of all potential swabbing surfaces in the facility. Five of these surfaces were swabbed every day during the study and were considered the 5 greatest-risk sentinel surfaces. Ten surfaces were swabbed daily and were rotated among the remaining locations and were considered systematic surfaces. Sampling occurred near the end of work shifts and before surfaces were cleaned and disinfected. Four swabs (Transport Swab 5-42, Technical Service Consultants, Heywood, Lancaster, UK for Europe locations or Enviro Swabs, 3M, St. Paul, Minnesota, USA for USA locations) were used for each sampling surface, with each individual swab used to sample a 25 cm^2^ area. The next three swabs were used to swab the same surface adjacent to the first swabbing area, also sampling a 25 cm^2^ area, and so on. For smaller surfaces that did not have enough surface area to accommodate 4 adjacent swabbings, the sampling individual could select 4 similar items nearby to sample, such as 4 sink nobs or 4 elevator buttons in the same location. Site managers were encouraged to sample all areas of the facility over the 14-day study period.

At the St. Cloud location, the site manager created a sampling plan focused on direct- touch items by the SARS-CoV-2 infected resident and high-frequency touch point items by facility staff. The purpose of the sampling plan was to confirm that the facility hygienic practices were effective in preventing spread of SARS-CoV-2 beyond immediate touch surfaces of the infected resident.

After collection, labeled swabs were sent via overnight courier to Eurofins Steins Laboratorium, Vejen, Denmark for Europe locations or Eurofins Microbiology Laboratories, Battle Creek, Michigan, USA for USA locations and tested using a multiplex RT-PCR assay for Coronavirus Envelop E gene (VIR*Seek* Screen, Eurofins GeneScan, Freiberg, Germany) and SARS-CoV-2 RdRP gene (VIR*Seek* Ident, Eurofins GeneScan). Coronavirus PCR reactions with C_q_ values ≤38 were considered detected, with LOD of 7 copies per reaction. SARS-CoV-2 reactions with C_q_ values ≤39 were considered detected, with LOD of 19 copies per reaction.

## Results & Discussion

### Clinical Testing

In two rounds of volunteer employee RT-PCR clinical testing, three volunteers tested positive for SARS-CoV-2 on week 1, two in Saverne (C_t_ 33 and 36) and one in New Orleans (C_t_ 35). Two volunteers tested positive in week 2 testing, one in Saverne (C_t_ 35) and one in Milan (C_t_ 33) (Table 2). None of the employees were displaying symptoms of COVID-19 at the time of testing. Following host company policies, the positive employees were immediately placed on administrative COVID-19 leave when laboratory results were received (two days after sample collection). None of these employees developed COVID-19 symptoms during their respective quarantine periods. No other study locations had RT-PCR clinical evidence of active COVID19 infection among their workforce. Although a majority of on-site employees were tested at each study location, as only volunteers were tested a number were not, which raises the question of whether there were other employees potentially able to shed virus into the workplace during the study.

**Table 2.**
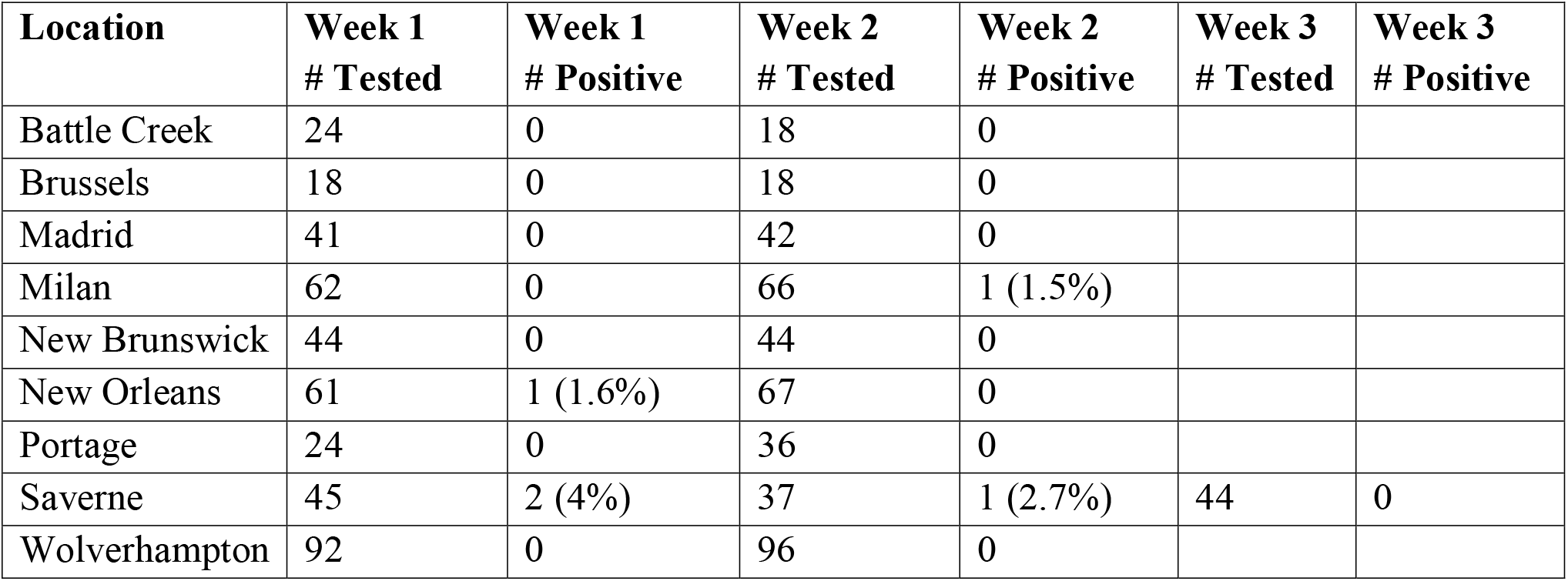
Study location employee volunteer SARS-CoV-2 detection rates.

| **Location** | **Week 1**<br>**# Tested** | **Week 1**<br>**# Positive** | **Week 2**<br>**# Tested** | **Week 2**<br>**# Positive** | **Week 3**<br>**# Tested** | **Week 3**<br>**# Positive** |
| --- | --- | --- | --- | --- | --- | --- |
| Battle Creek | 24 | 0 | 18 | 0 |  |  |
| Brussels | 18 | 0 | 18 | 0 |  |  |
| Madrid | 41 | 0 | 42 | 0 |  |  |
| Milan | 62 | 0 | 66 | 1 (1.5%) |  |  |
| New Brunswick | 44 | 0 | 44 | 0 |  |  |
| New Orleans | 61 | 1 (1.6%) | 67 | 0 |  |  |
| Portage | 24 | 0 | 36 | 0 |  |  |
| Saverne | 45 | 2 (4%) | 37 | 1 (2.7%) | 44 | 0 |
| Wolverhampton | 92 | 0 | 96 | 0 |  |  |

### Environmental Surface Testing

At the New Orleans study location, pre-study baseline environmental sampling revealed a 4% Coronavirus prevalence rate on 48 surfaces using a single swab per surface. On the day after the first day of clinical testing, Coronavirus detection rate was 13% of 60 swabs on 15 surfaces. The greater rate of surface detections corresponded with detection of a clinically positive employee. The daily number of positive swabs at the study location declined as the study progressed (Figure 1), which suggests that removing the infected employee from the workplace resulted in reduced number of contaminated environmental surfaces. The pre-study baseline testing positives (log book and photocopier) may have been leading indicators of the presence of a preclinical or asymptomatic SARS-CoV-2 employee; these two locations were frequented by this employee. Furthermore, after notification of the employee positive test result, that employee’s immediate work area (desk, computer mouse, keyboard, and chair) was swabbed two days after the employee was at the location, and none of the touch-point surfaces were positive for Coronavirus. That said, a number of high-frequency-touch points surfaces were found to be contaminated several days after the clinical testing was done and for several days afterward. Several of these surfaces had been touched by the infected employee (elevator buttons, break room chairs, break room door handle). Not all employees at this study location volunteered for clinical testing so others also may have contaminated these locations. The infected employee was not displaying symptoms of COVID-19.

**Figure 1.**
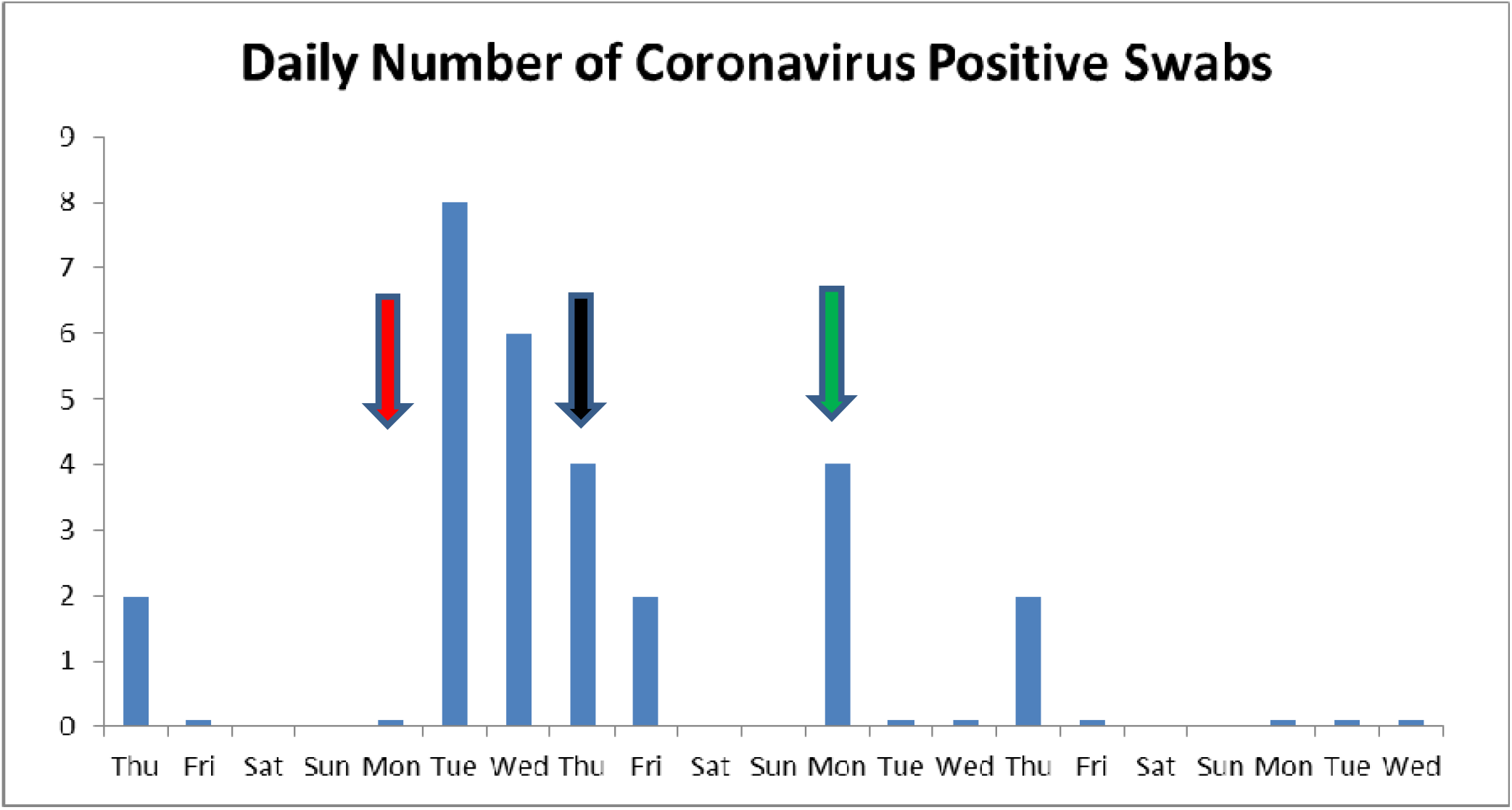
New Orleans daily number of Eurofins COVID-19 Sentinel^™^ Coronavirus positive environmental surfaces. Arrows indicate days when clinical samples for SARS-CoV-2 were collected (green arrow = no employees positive; red arrow = employee positive) and when clinically positive employee was excused from workplace (black arrow). Samples were not collected on weekend days.

At the Saverne study location (Figure 2), a work area that one of the three infected employees worked at was found to be contaminated with Coronavirus (3 work bench positive swabs). These Coronavirus-positive locations were confirmed as SARS-CoV-2. Unlike the New Orleans location, common high-frequency touch point areas were not contaminated with Coronavirus. The Milan location had one external door handle Coronavirus positive on day 14. Together, these findings suggest that workplace surface contamination was variable at locations where clinically positive employees were present. The data also demonstrate the importance of using sample volumes large enough to cover the number of high-frequency-touch point surfaces in a facility. And, like the New Orleans study location, both Saverne and Milan locations were able to resolve Coronavirus environmental contamination with attentive disinfection practices and SARS-CoV-2 positive employee quarantines.

**Figure 2.**
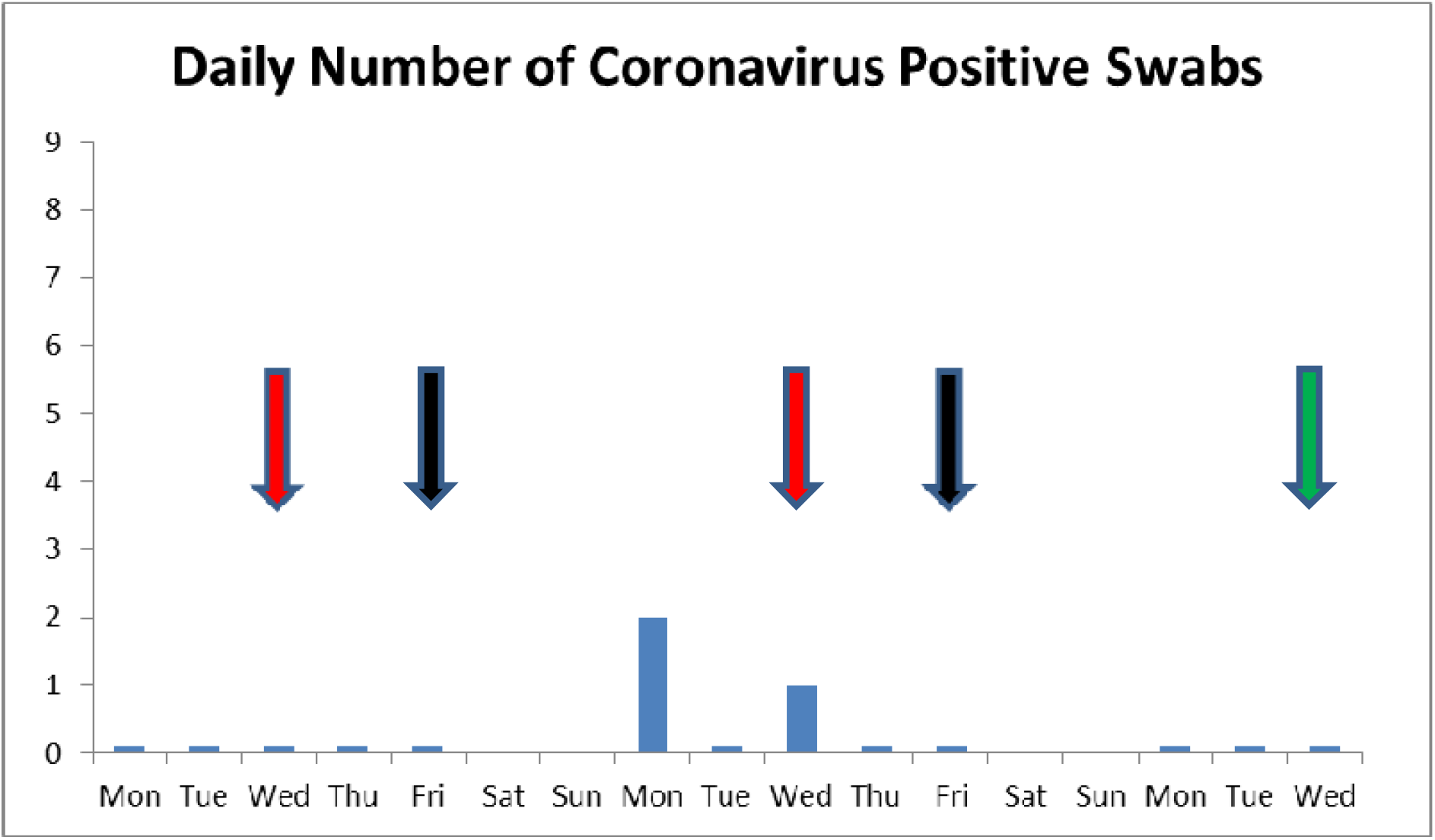
Saverne daily number of Eurofins COVID-19 Sentinel^™^ Coronavirus positive environmental surfaces. Arrows indicate days when clinical samples for SARS-CoV-2 were collected (green arrow = no employees positive; red arrow = employee positive) and when clinically positive employees were excused from workplace (black arrow). Samples were not collected on weekend days.

At most other study locations with no clinically identified SARS-CoV-2 positive employees, very few or no Coronavirus environmental surface detections were found (14% of total positive surfaces). The Madrid location had two Coronavirus positive surfaces on a package reception table and an outer entrance push button. The Battle Creek and Portage locations each had one Coronavirus positive surface location (plastic bin and keyboard, respectively). The few positive surface detections at these locations may be due to the presence of infected workers that were not tested or COVID positive non-employee vendors, delivery drivers, or custodians. The New Brunswick study location was the notable exception. At this site, seven office-related locations were Coronavirus positive (file drawers, paper hole punch, keyboard, toilet handle, equipment keypads) on the first day of sampling. No clinical SARS-CoV-2 employees were identified by RT-PCR testing at either sampling time; however, on the second round of testing, employees also were administered blood draws for SARS-CoV-2 antibody testing conducted by Viracor Eurofins. The antibody testing was done outside the present study protocol. Five employees were positive for SARS-CoV-2 IgM and one employee was positive for SARS-CoV- 2 IgG (a total 14% antibody prevalence rate among tested employees). All six of these employees tested negative for SARS-CoV-2 by RT-PCR testing. Prior to commencement of the study, the IgG positive employee was mildly symptomatic while living with a COVID-19 diagnosed individual. This employee recently returned to work after a 14-day quarantine period. RT-PCR may not always detect infected individuals due to sampling irregularity, timing of sample collection, limits of detection, or other factors (*15*). It is reasonable to surmise that the positive Coronavirus environmental surface samples may have predicted the presence of SARS-CoV-2 exposed employees.

Odds ratio calculation for locations with clinical PCR or Antibody positives (2,400 environmental swabs) vs. locations without clinical positives (3,000 environmental swabs) reveals that locations with Coronavirus positive environmental surfaces had 10 times greater odds (95% confidence, P=0.000001) of having clinically positive employees compared to locations with no positive surfaces.

The VIR*Seek* Screen Coronavirus E-gene test will detect other acute respiratory Coronaviruses in addition to SARS-CoV-2; therefore, it is possible that some detections reported here may be due non-COVID-19 causing strains. However, the prevalence of circulating seasonal Coronavirus infections is low (0.6 to 2.2%) as demonstrated in a large study of over 800,000 samples collected in the USA (*16*). SARS-CoV-2 population infection prevalence rates also are assumed to be low except in communities with active outbreak transmission. For example, a 5% SARS-CoV-2 prevalence rate was seen in patients with mild influenza-like illness in a Los Angeles, California medical facility (*17*). In an ongoing New Jersey study, a 4% SARS- CoV-2 antibody prevalence rate was reported among 629 households and 1,062 volunteer individuals participating in a community surveillance program (*18*). The employee antibody prevalence rate at the present New Brunswick New Jersey study location was 3.5 times greater than the New Jersey community surveillance rate. This implies a greater SARS-CoV-2 exposure history among the location employee corps compared to nearby communities. It is not surprising then that Coronavirus positive environmental surfaces were found at this location, but only at the beginning of the study. Perhaps this was due to latent virus shedding by an employee who resolved the infection in their oropharyngeal area by the time the study was initiated and was no longer shedding a detectable virus titer as the study progressed.

For all environmentally positive locations it is interesting to note that a number of surfaces (break room chairs, break room door handles, work benches, and water faucet handles) were repeatedly tested positive for Coronavirus during the sampling period (Table 2). At the New Orleans location, a heat map showing a cluster of Coronavirus contaminated environmental surfaces reveals proximity of elevators and break room surfaces (entryway door and chairs) that were repeatedly tested positive (Figure 3). The repeat positives are suggestive that virus RNA may persist on surfaces for several days, which agrees with previous reports (*3, 19*) or may suggest that an undiscovered untested or newly infected employee remained in the workforce during the study. Indeed, some study locations had several on-site employees that did not volunteer for clinical testing, some of whom may have been active shedders of SARS-CoV-2. In addition, non-employee individuals had access to the facilities, including vendors and custodian personnel. These repeat-positives surfaces may merit extra consideration as useful sentinel monitoring sites.

**Figure 3.**
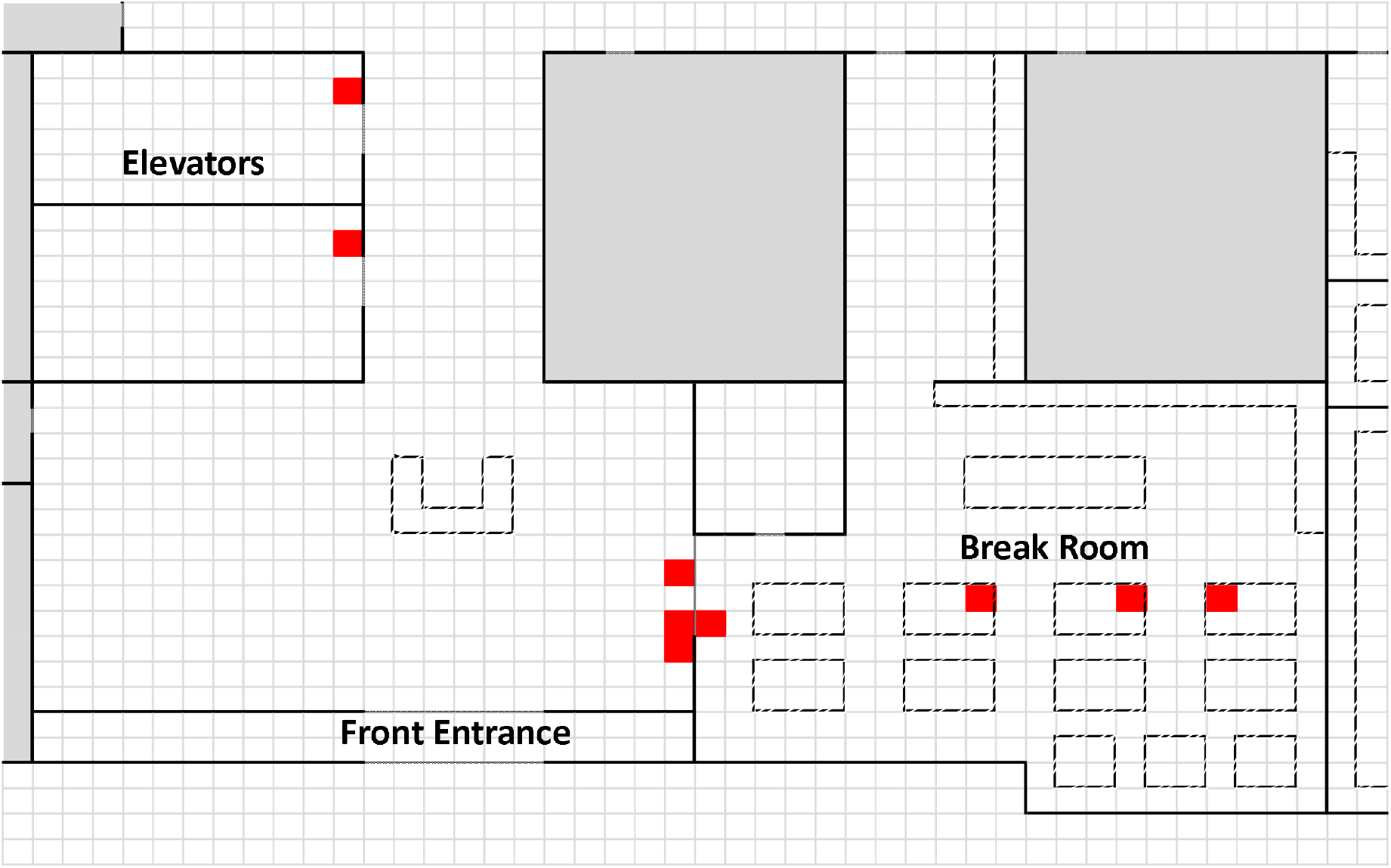
New Orleans location cluster heat map of positive Coronavirus environmental surfaces in break room and elevators. The main building entrance is shown at bottom left.

Despite repeated simultaneous adjacent sampling (4 swabs per surface per day) of individual surfaces, few surface locations (elevator buttons, computer mouse, and twice with a break room chair) had more than one swab per day show Coronavirus detection, with 2 of 4 swabs positive. All other surface detections occurred with only 1 of 4 swabs positive. It was never observed that all four samples were positive. When virus RNA was detected, RT-PCR C_q_ values suggests Coronavirus RNA titer was small (range 34 to 38 cycles, average C_q_ of 36 for 44 detections) and near the limit of detection of the method (Table 3). These C_q_ results were similar to those reported in COVID-19 patient hospital rooms before disinfection (*11*). These findings suggest that even in locations in close proximity to an active virus shedder, viral RNA loads on environmental surfaces were small and not uniformly distributed on a surface.

**Table 3.** High-frequency-touch points testing positive for Coronavirus E gene and associated RT-PCR cycle quantification (C_q_) values.

| **Touch Point** | **Number of Detections** | **$C_q$** |
| --- | --- | --- |
| Break room chair | 6 | 35, 35, 35, 36, 36, 37 |
| Break room door handle | 6 | 34, 34, 35, 36, 36, 38 |
| Work bench surface | 5 | 36, 36, 37, 38, 38 |
| Equipment control panel | 2 | 35, 36 |
| Photocopier control panel | 2 | 36, 37 |
| Log book | 2 | 35, 36 |
| Men's toilet faucet handle | 2 | 35, 35 |
| Women's toilet faucet handle | 2 | 35, 36 |
| Elevator buttons | 2 | 34, 36 |
| Computer mouse | 2 | 35, 36 |
| Keyboard | 2 | 36, 36 |
| File drawer handle | 2 | 35, 36 |
| Entrance push button | 1 | 38 |
| Work bench hot water faucet handle | 1 | 35 |
| Workstation refrigerator door handle | 1 | 35 |
| Office desk | 1 | 36 |
| Stock room exit door handle | 1 | 35 |
| Workspace exit door handle | 1 | 36 |
| Furnace oven door handle | 1 | 34 |
| Plastic bin | 1 | 37 |
| Office paper hole punch | 1 | 36 |

Managers at the study locations with recurrent surface positives responded by assessing the effectiveness of disinfection controls at the site. For example, the landlord of the New Orleans facility provided building cleaning service. This service used quaternary ammonium compound disinfectants on the Environmental Protection Agency List N (EPA Registration Numbers 10324-157-12502 and 1839-169-12502). The cleaning service practice was to spray and wipe most high-frequency-touch point surfaces with the disinfectants, while keyboards, computer mice, and computer screens were dusted rather than disinfected. Two deficiencies were noted with this practice. First, contact time after disinfectant application to surfaces should be at least 10 minutes before wiping. Second, a wiping cloth was repeatedly reused rather than single-use paper towels. At the New Orleans location, exclusion of the SARS-CoV-2 infected employee and improvements in sanitation practices (use of disposable towels and longer disinfectant contact time) likely contributed to the decline in number of positive swabs shown in Figure 1.

At the St. Cloud study location, three Coronavirus-contaminated surfaces were found. Two nurse call buttons (C_q_ 35 and 35) and one from a wheelchair arm (C_q_ 34). Two of the three samples (nurse call button and wheelchair arm from the same resident room) were confirmed as SARS-CoV-2 with C_q_ values of 37 and 38. Samples obtained within the resident’s room including bed frame, door handles, air intake vents, sink faucet handle, and the nightstand were all negative. Samples outside the resident’s room, including door handles to a soiled laundry room and exterior doors, were also negative. Prior to having its first resident diagnosed with COVID-19, the organization created a contingency plan for segregating diagnosed residents into a re-designed multi-room section of the facility. The redesign included erecting physical separation (temporary walls), creating negative pressure in the residential rooms, serving in-room meals to the residents, and having staff dedicated to the area. Results of the environmental surface testing confirmed that these COVID-19 controls were working.

## Conclusion

Study locations with no evidence of clinical employee COVID-19 infections had no to very few Coronavirus surface detections. This suggests that non-SARS-CoV-2 Coronavirus strains were not widely circulating in the employee populations at the study locations. At locations where SARS-CoV-2 positive employees were detected, there was a strong association with finding contaminated high-frequency-touch point surfaces. All employees who tested positive were asymptomatic and passed daily fit for duty screens before and two days after testing. Therefore, very frequent testing of staff would appear to be needed to detect asymptomatic and presymptomatic employees. Locations with numerous environmental positive detections for Coronavirus all had a clinically positive employee during the study period; therefore, environmental testing results appear to be a useful tool to inform clinical testing need in order to detect asymptomatic or presymptomatic virus spreaders. Workplaces with surface coronavirus detections responded with interventions that reduced detection prevalence rates. This study provides evidence that Coronavirus environmental monitoring can be a useful tool to help manage COVID-19 incidence in the workforce.

## Data Availability

Data is available from the corresponding author.

## Acknowledgement

Funding for this study was generously provided by the participating locations and participating laboratories.

## Notes

### Competing Interest Statement

The authors have declared no competing interest.

### Clinical Trial

David C. Drovetta, Director of Regulatory Affairs and Quality Assurance at Viracor Eurofins, has a written letter (see supplemental file) stating that the study design has met their ethics and privacy policy, including signed informed consent of all study volunteers and acknowledgement that results will be submitted for publication. Volunteers were tested for SARS-CoV-2 and the study did not include a clinical trail.

### Funding Statement

Funding for this study was provided by the participating locations and participating laboratories. No authors or institutions received payment or services from a third party.

